# Disrupted microbial queuosine biosynthesis contributes to dysbiosis in human inflammatory bowel disease

**DOI:** 10.64898/2026.10.12.26365231

**Authors:** Yueqing An, Yinglin Xia, Jun Sun

## Abstract

**Background and Aims:** As a microbiome/diet-derived vitamin, queuine (q) is a micronutrient that eukaryotes acquire from microbiota or diet. Vitamin q, the free base form of queuosine (Q), is used for Q-modification of transfer RNA (tRNA). Although tRNA-Q-modification in regulating eukaryotic cell physiology is recognized, the relationship between Q-tRNA-modification and the human microbiome in health and inflammatory bowel diseases (IBD) remains unknown. Which bacteria produce q and how the q-producing bacteria contributing to human health and IBD are not explored. Here, we hypothesize that disrupted microbial Q-biosynthesis contributes to the dysbiosis of human IBD. We performed comprehensive multi-omics integrative analyses using publicly available datasets. Specifically, we re-analyzed publicly available shotgun metagenomic, single-cell RNA sequencing (scRNA-seq), metabolic datasets, and completed meta-analysis.

**Methods:** To systematically investigate microbial Q-biosynthesis in healthy individuals and IBD patients, we performed a systematic secondary analysis by integrating 9 publicly available metagenomic datasets, 15 scRNA-seq datasets and one metabolic dataset published prior to 2026. We present a novel secondary bioinformatic reanalysis of sequence outputs mapped to the latest microbial taxonomy, functional pathways and microbial Q-related genes. Longitudinal study of shotgun metagenomics across the human lifespan (3146 samples) enabled taxonomic and functional profiling.

**Results:** We first identified key q-producing bacteria (e.g., *Bacteroides* and *Alistipes*) in healthy populations. We then found that patients with IBD had significantly reducation of q-producing bacteria and a coordinated downregulation of the Q-related pathways, including Q, preQ_0_, de novo guanosine ribonucleotide biosynthesis and de novo guanosine deoxyribonucleotide biosynthesis pathways. This functional impairment was primarily driven by reduced contributions from *Bacteroides*, *Alistipes and Akkermansia*. The longitudinal stability of the q-producing bacteria community across the human lifespan (1638 samples). We observed a significant negative correlation between host age and the abundance of q-producing bacteria in patients with UC. Moreover, Queuine tRNA-ribosyltransferase 1 (QTRT1) expression in human intestinal epithelial cells was significantly reduced in patients with IBD compared to healthy controls.

**Conclusions:** Our study used multi-omics datasets to define a novel functional dysbiosis characterized by a reduction of microbial Q-biosynthesis in patients with IBD. Vitamin q and its tRNA modification is lesser-recognized in human IBD. Understanding the cellular and organismal mechanisms of this microbiome-dependent micronutrient will advance the prevention of IBD and improve the quality of life of patients with IBD.

**What is already known on this topic?:** Queuine, vitamin q, is a highly conserved hypermodified nucleoside micronutrition could only be acquired from microbiota or diet. It is used for modification of transfer RNA to protein transitional fidelity, but the specific microbial pathways evolved and the relationship with IBD are unclear.

**What this study adds:** This study identifies a novel functional dysbiosis in IBD patients characterized by the depletion of q-producing gut bacteria (e.g., *Bacteroides* and *Alistipes*) and the downregulated microbial Q biosynthesis pathways, which correlates with host QTRT1 deficiency to potentially drive IBD pathogenesis.

**How this study might affect research, practice or policy:**

1. 1. Identifies the altered Q-biosynthesis pathway in human IBD. Establishes the “Microbial Q-synthesis-Host tRNA modification” axis as a novel mechanistic link, shifting human IBD microbiome research from taxonomic description to functional epitranscriptomics.
2. Overcomes the inherent biases of traditional meta-analyses through a unified secondary bioinformatic reanalysis, leveraging HMP unified metabolic analysis network 3 (HUMAnN3)-based modeling of 3,000+ samples, to reveal a universal breakdown of the microbial Q-biosynthetic supply chain across all IBD subtypes.
3. Recognize vitamin q as a critical gut-health micronutrient, influencing dietary guidelines and the regulation of functional medical foods for IBD management.

## Introduction

Vitamin q, is an essential and less-recognized micronutrient that eukaryotes cannot synthesize de novo but must acquire from microbiota or dietary sources. In eukaryotes, q is used for Q-tRNA-modification that governs protein translational fidelity and efficiency ^1^. Vitamin q is incorporated into tRNAs with GUN anticodons (asparagine, aspartic acid, histidine, and tyrosine) by the host enzyme complex Queuine tRNA-ribosyltransferase 1 and 2 (QTRT1/QTRT2) ^2–4^. Disruption of tRNA-modifications has been increasingly linked to human diseases^5,6^, including neurodegenerative disorders^7^, cancer^8–11^, diabetes^6,12,13^, and mitochondrial diseases^14,15^. Our previous study has demonstrated that Q-tRNA-modifications play an unexplored novel role in the pathogenesis of intestinal inflammation by altering epithelial proliferation and junction formation^16^. Because q can’t be synthesized de novo in human cells, the host is entirely dependent on gut bacteria and/or diets for essential Q-tRNA-modification, However, the status of q-producing bacteria in health and IBD is still unknow. This critical microbial-host link in chronic intestinal inflammation remains largely unexplored.

In the context of the human gut, the microbial metabolic capacity is profoundly altered in patients with IBD. Beyond mere taxonomic shifts, a state of functional dysbiosis emerges, characterized by the loss of specific microbial outputs. IBD, comprising Crohn’s disease (CD) and Ulcerative colitis (UC), represents a complex breakdown of the homeostatic dialogue between host genetics, the gut microbiome, and dietary exposures, which imposes a profound biological and socioeconomic burden, evidenced by a significant reduction in life expectancy of five to eight years^17,18^. It is known that q becomes essential for survival in germ-free mice under tyrosine-deficient conditions, revealing a critical link between the q-axis and tyrosine metabolism^19^. Although Q-biosynthesis is a strictly bacterial trait, it is not universally produced by all bacteria. Comparative genomics distinguishes between q-producer, who is capable of de novo synthesis and q-consume, who dependent on the salvage of Q or its intermediates^20,21^. We thus hypothesized that disrupted microbial Q-biosynthesis contributes to the dysbiosis of human IBD.

To systematically investigate microbial queuosine biosynthesis in healthy individuals and patients with IBD, we performed comprehensive multi-omics integrative analyses using publicly available datasets. Specifically, we aimed to (1) identify q-associated bacterial taxa depleted in IBD through a meta-analysis of nine public gut metagenomic datasets, (2) quantify the genomic abundance of bacterial Q-biosynthesis functional pathways (including Q, preQ_0_, de novo guanosine ribonucleotide biosynthesis and de novo guanosine deoxyribonucleotide biosynthesis pathways) by profiling shotgun metagenomic data from publicly available datasets, and (3) assess host QTRT1 expression in IBD versus healthy controls using scRNA-seq data. We present a novel secondary bioinformatic reanalysis of sequence outputs mapped to the latest microbial taxonomy. While previous studies have largely focused on descriptive compositional changes in the gut microbiota, they often fail to pinpoint the exact microbial contributors to specific functional outputs. To bridge this gap, we employed a novel, high-resolution integrative framework HUMAnN3^22^. Especially, we used a linear mixed-effects model (LMM)-based meta-analysis and model selection to account for the study heterogeneity and to adjust for the age and gender effects. This approach represents a significant methodological leap, unlike traditional tools that provide a bacteria taxa and genes overview, HUMAnN3 allows us to identify not only the Q-biosynthesis capacity depleted in IBD, but also which microbial species responsible for this functional loss. Identifing the Q-biosynthesis pathway will allow us to better understand the pathogenesis of human IBD, develop new diagnostic biomarkers. Furthermore, vitamin q supplementation or q-producing bacteria could be targets for new therapeutic strategies.

## Results

### Methodological framework resolves top 20 q-producing bacteria species and top 20 functional pathways in human gut

Human body acquires Vitamin q from intestinal microbiota or diet. To systematically investigate which bacteria produce q and how the q-producing bacteria contribute to human health and IBD, we have designed the studies by performing a comprehensive multi-omics integrative analysis using publicly available datasets. Altogether 64 datasets were sought for full-text screening, and 25 datasets from Google Scholar, PubMed, The Inflammatory Bowel Disease Multi’omics Database (IBDMDB), National Center for Biotechnology Information-Gene Expression Omnibus (NCBI-GEO) and European Molecular Biology Laboratory-European Bioinformatics Institute (EMBL-EBI) were included in our multi-omics analysis. By re-analyzing shotgun metagenomic and scRNA-seq datasets, our study aims to define a novel functional framework of in health and disease pathogenesis (**Fig. 1A**). We first identified the top 20 q-producing bacterial species based on their average relative abundance across healthy gut. The taxonomic landscape was overwhelmingly dominated by beneficial obligate anaerobes, with *Bacteroides vulgatus* exhibiting the highest baseline abundance, followed by core commensals including *Bacteroides massiliensis*, *Akkermansia muciniphila* and *Bacteroides uniformis* (**Fig. 1B**). To establish a functional baseline for Q-process, we clarify them by further analyzing the key enzymatic steps, mapping related biosynthesis pathways including the Q-biosynthesis pathway and guanosine ribonucleotide de novo biosynthesis, which is the conversion of Guanosine triphosphate (GTP) to preQ_0_ and the final insertion of Q into tRNA by tRNA-guanine transglycosylase (TGT), across the identified biosynthesis pathways (**Fig. 1C**).

**Figure 1.**
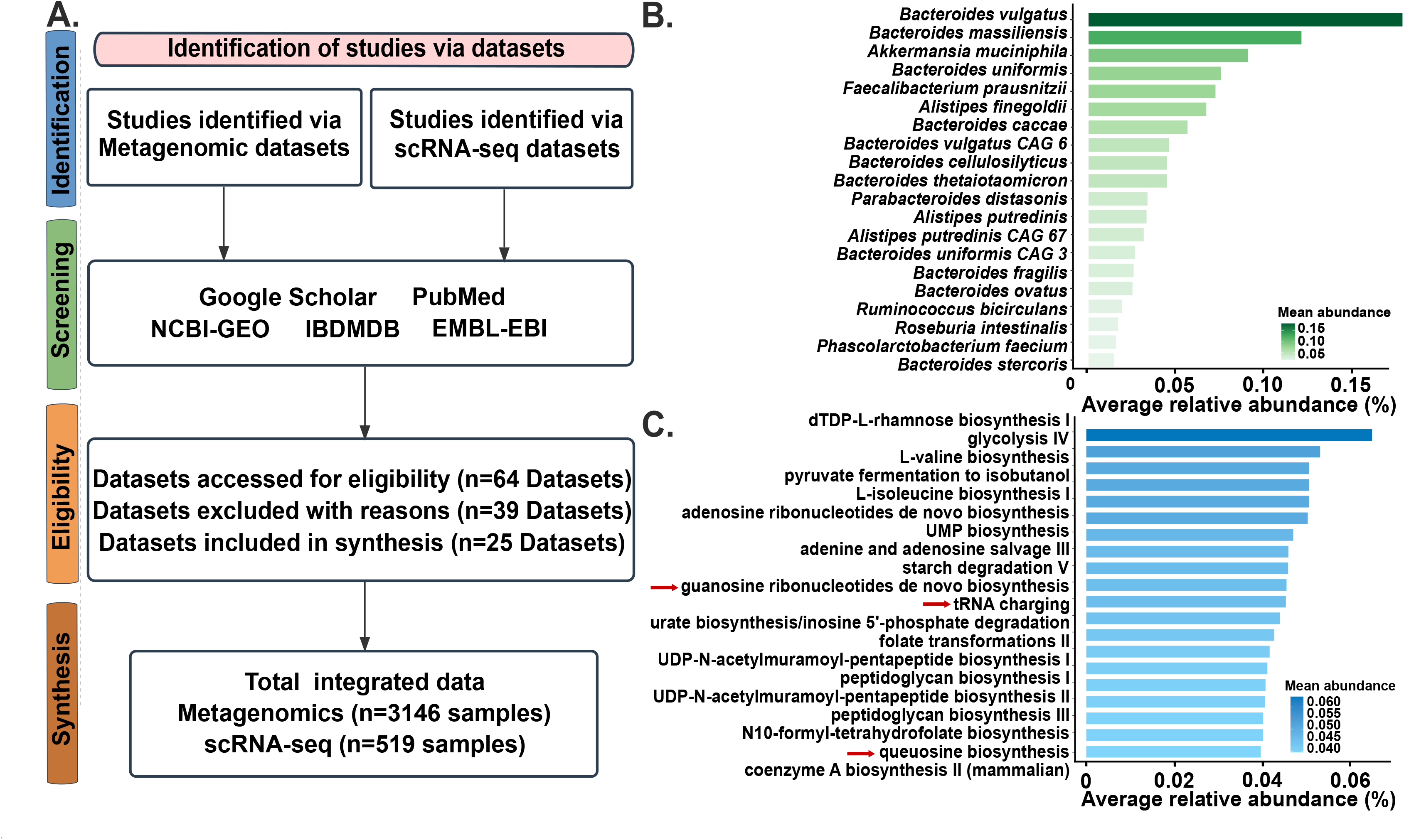
Methodological framework and compositional landscapes of top 20 bacteria and top 20 functional pathways in the healthy control human gut. **(A)** Preferred reporting items for systematic reviews and meta-analyses including metagenomics and scRNA-seq datasets define the functional role of Q-tRNA-modification in human IBD (CD and UC). **(B)** Taxonomic profile highlighting the top 20 bacterial species within the healthy population. Species are ranked in descending order based on their mean relative abundance across the pooled healthy control cohorts, led by core intestinal commensals including *B. vulgatus*, *B. massiliensis*, *A. muciniphila* and *B. uniformis*. Unclassified taxa were excluded prior to ranking, n = 429 healthy control samples. **(C)** Top 20 metagenomic pathways in healthy control samples including the Q-biosynthesis pathway and de novo guanosine ribonucleotide (red arrows). Unmapped and unintegrated pathways were excluded prior to ranking, n = 429 healthy control samples.

### Reduction of q-producing bacteria *Bacteroides* and *Alistipes* species across CD and UC

To establish a taxonomic baseline for further Q-processing analysis in the human gut, we investigated bacteria distribution and modification at the species level and observed a significant downregulation of key q-producing bacteria, e.g., *Bacteroides* and *Alistipes* in both CD and UC patients, compared with healthy controls by analyzing the taxonomic profiles. Numerous species within the *Bacteroides* genus (e.g., *B. massiliensis*, *B. plebeius* in both CD and UC, *B. eggerthii* in CD) (**Fig. 2A, C)** and *Alistipes* genus (e.g., *A. putredinis, A. shahii* in both CD and UC *A. senegalensis* in UC) were significantly reduced (**Fig. 2B, D**).

**Figure 2.**
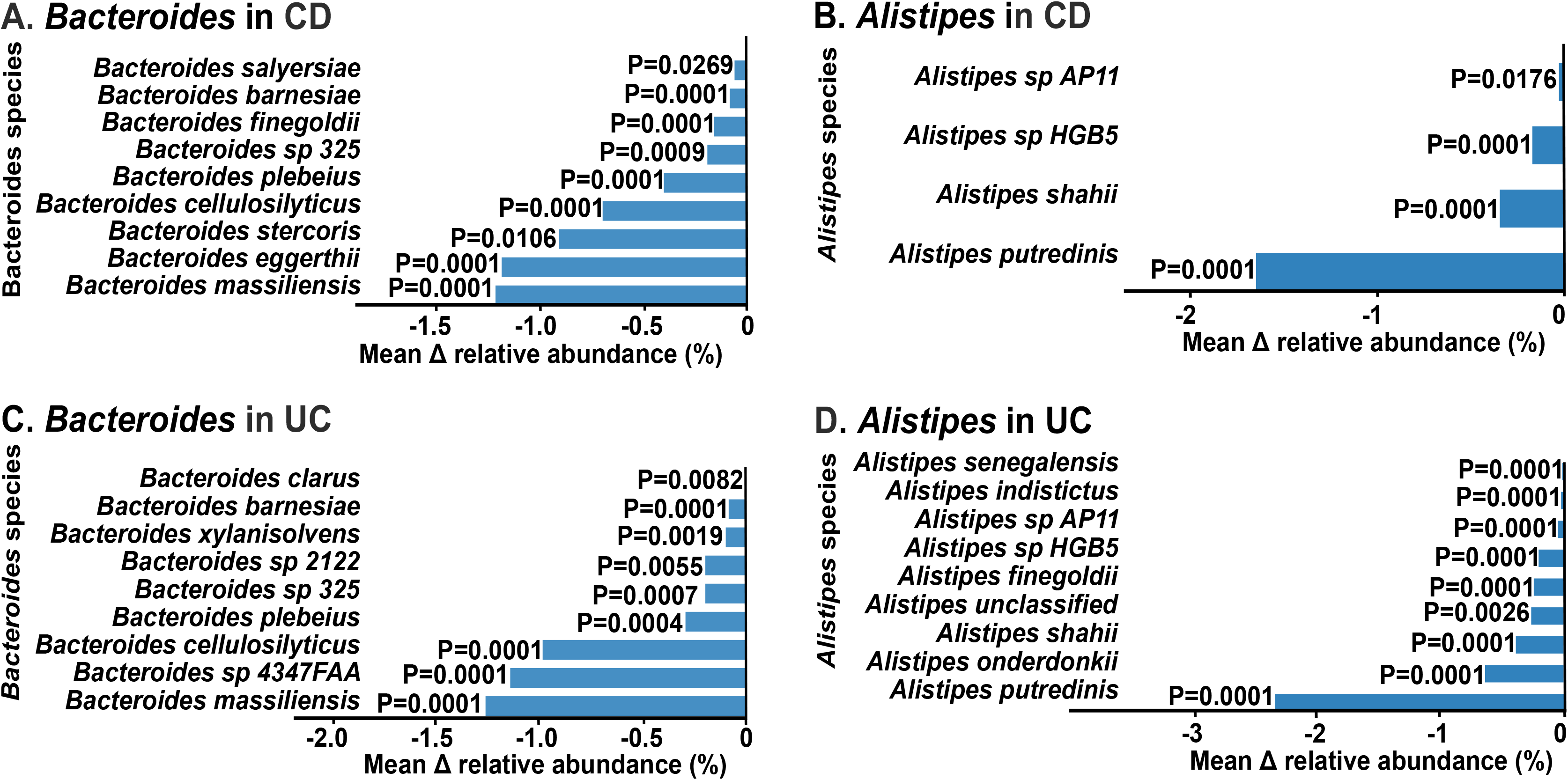
The synchronized downregulation of q-producing bacteria *Bacteroides* and *Alistipes* in both CD and UC compared to healthy controls. **(A)** *Bacteroides* and **(B)** *Alistipes* species that are significantly reduced in CD patients compared with healthy controls. Species-level reduction of significantly altered Q-producers **(C)** *Bacteroides* and **(D)** *Alistipes* in UC patients. Data are expressed as mean difference in relative abundance, Wilcoxon rank sum test, n=3146 samples (1406 CD, 830 UC and 910 healthy controls).

### Q-process pathways shift: decrease of *Bacteroides* and *Alistipes* across both CD and UC

Nine studies were included in the qualitative synthesis, of which eight (1406 samples) contributed data on CD studies, six (830 samples) contributed data on UC studies and nine (910 samples) on healthy control studies. Metagenomic analysis showed relative contribution of bacteria species to Q-related pathways, including the Q-biosynthesis pathway, preQ_0_ biosynthesis pathway, de novo guanosine ribonucleotide biosynthesis pathway, and de novo guanosine deoxyribonucleotide biosynthesis pathway (**Fig. 3, 4, S1, S2**). Schematic diagram of the microbial Q-associated biosynthesis pathways are listet (**Fig. 3A**). Starting from GTP, sequential enzymatic reactions via preQ_0_ and preQ_1_ lead to the formation of Q-tRNA, illustrating the key metabolic steps and regulatory precursors involved in microbial Q modification. Specifically, the genomic abundance of the direct Q-biosynthesis pathways including the Q-biosynthesis pathway and preQ_0_ biosynthesis pathway contributed by *Bacteroides* spp. including *B. massiliensis* **(Fig. 3B, 3E)**, *B. eggerthii* **(Fig. 3C, 3F)**, *B. plebeius* **(Fig. 3D, 3G)***, B. intestinihominis* **(Fig. S1A, S1E)**, and *B. dorei* **(Fig. S1B)** showed a significant overall decrease in CD patients compared to healthy controls. While the guanosine related pathways, upstream pathways of Q-tRNA-modification, de novo guanosine ribonucleotide biosynthesis by the *Alistipes putredinis* exhibited depletion (**Fig. S1F**). Specifically, *Escherichia coli* demonstrated a significantly higher abundance in CD patients across the combined cohort in preQ_0_ biosynthesis pathway, Q biosynthesis pathway and de novo guanosine deoxyribonucleotide biosynthesis pathway (>**Fig. S1C, S1D, S1G**). Similarly, for the preQ_0_ biosynthesis and Q-biosynthesis pathways, the contributions of *B. dorei* **(Fig. 4A, 4D)**, *B. plebeius* **(Fig. 4B)***, B. massiliensis* **(Fig. 4E, S2A)**, *B. eggerthii* **(Fig. S2B, S2J)**, *B. xylanisolvens* **(Fig. S2C, S2E)***, B. intestinihominis* **(Fig. S2D)**, *and Akkermansia muciniphila* **(Fig. 4C, 4F)** were significantly lower in UC patients compared to healthy controls. Furthermore, the guanosine-related upstream pathways, which provide the essential precursors for Q-modification, showed a distinct loss of contribution from *Alistipes* spp. including *A. putredinis* **(Fig. S2F)***, A. senegalensis* **(Fig. S2G)***, A. indistinctus* **(Fig. S2H)** and *A. shahii* (**Fig. S2I**). The findings suggest that the functional impairment of microbial Q-biosynthesis in IBD is primarily driven by the specific depletion of key q-producing taxa, particularly within the *Bacteroides* and *Alistipes* genera.

**Figure 3.**
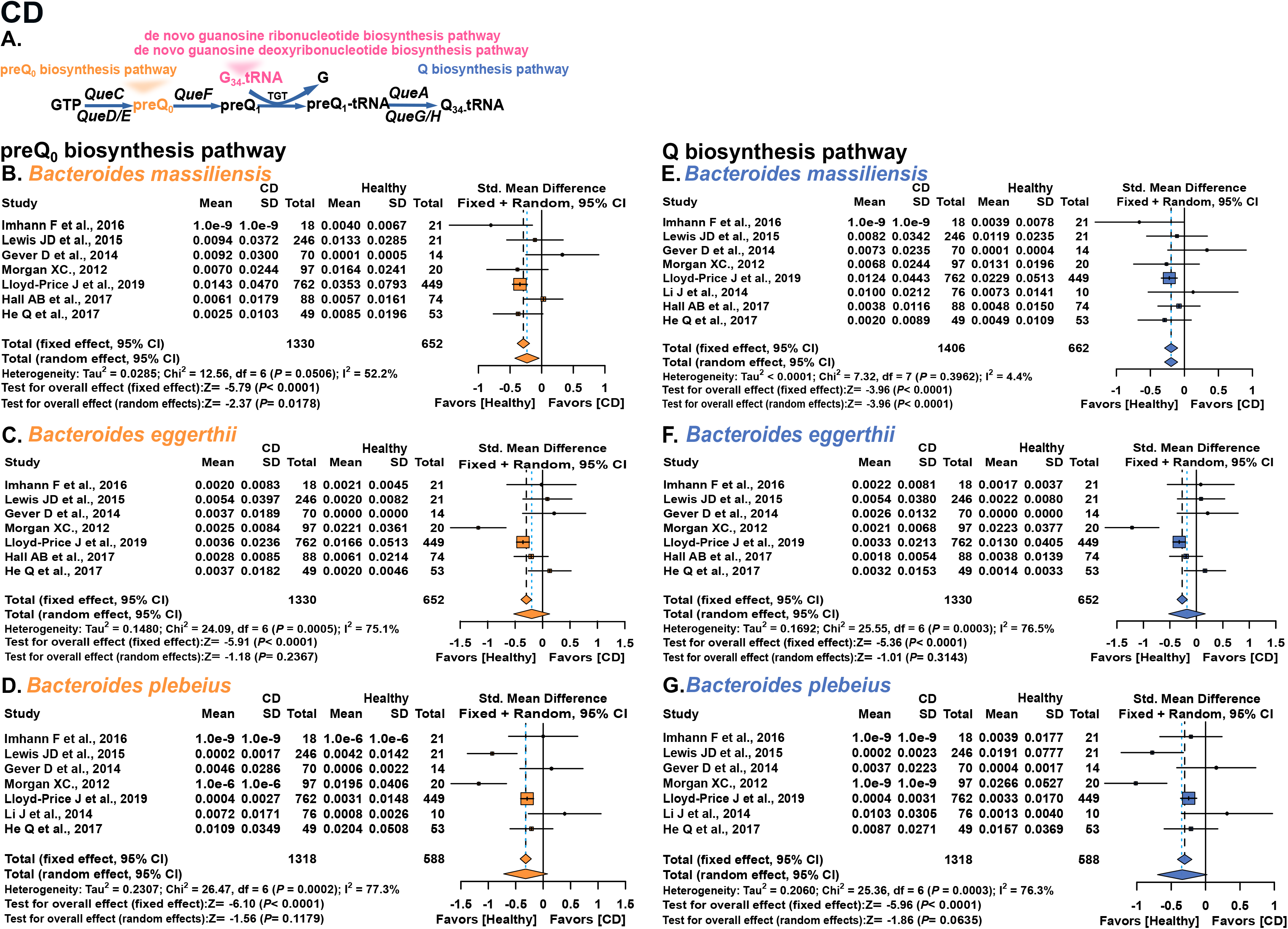
Divergent shifts in microbial Q-process pathways characterized by *Bacteroides* and *Alistipes* depression in CD. (**A**) Q-related biosynthetic pathways in bacteria and eukaryotes. Forest plots illustrate the Standardized Mean Difference (SMD) in the relative contribution for preQ_0_ biosynthesis pathway (highlighted in yellow) contributed by specific bacterial taxa (**B**) *B. massiliensis*, (**C**) *B. eggerthii*, (**D**) *B. plebeius*, and Q-biosynthesis pathway (highlighted in blue) contributed by (**E**) *B. massiliensis*, (**F**) *B. eggerthii*, (**G**) *B. plebeius* in CD patients. Squares represent the SMD for individual studies, with the size proportional to the study weight. Horizontal lines indicate the 95% CI. Diamonds represent the pooled effect size calculated via the random-effects and fixed-effects models. Heterogeneity is indicated by the I^2^ value and *P*-value provided at the bottom of each panel. Pooled *P*-values of both models are shown in each panel. Negative SMD values indicate depletion in CD patients, while positive values indicate enrichment. Each panel includes the studies with detectable contributions, n=2316 samples (1406 CD, 910 healthy controls).

**Figure 4.**
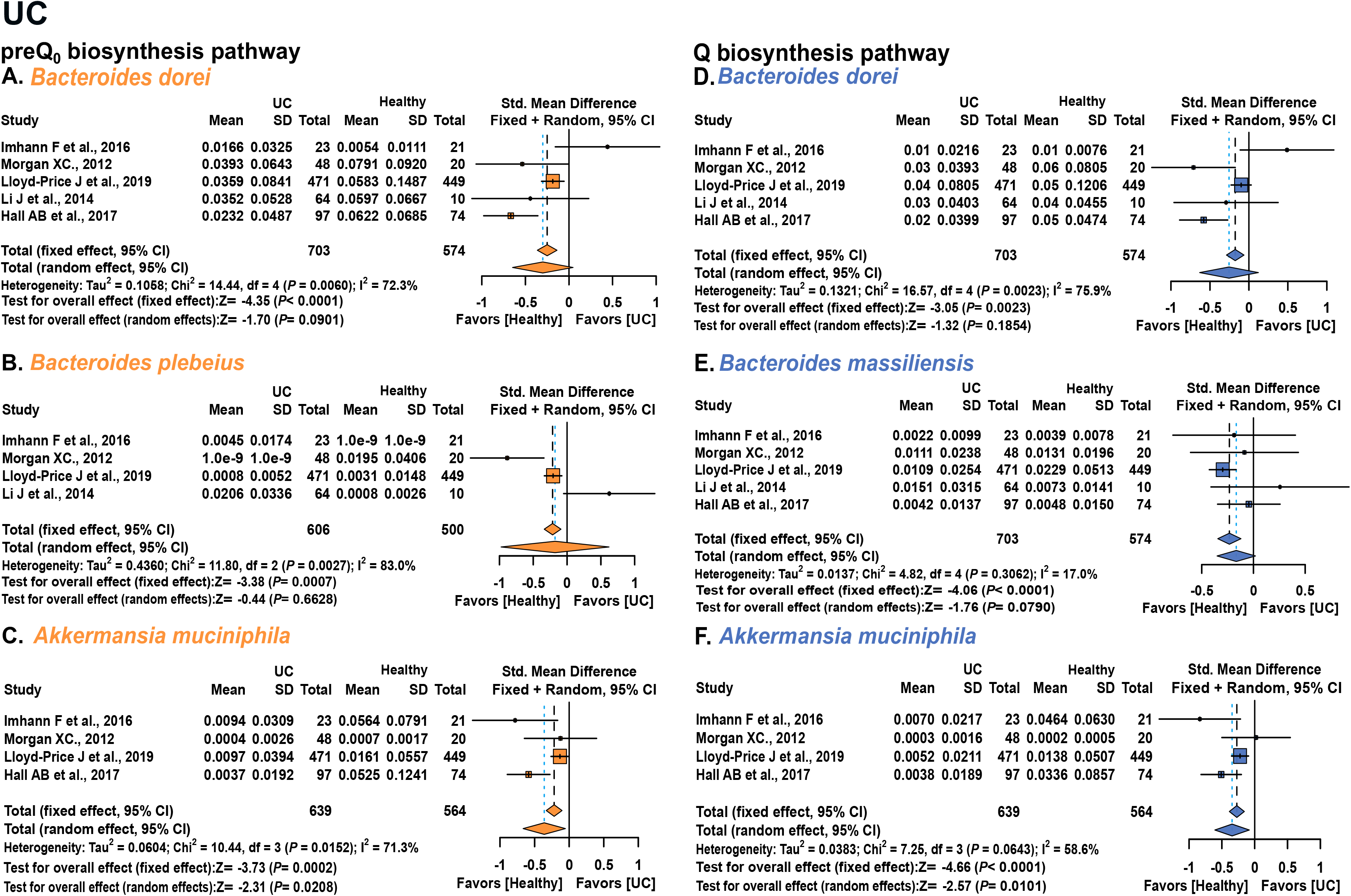
Decrease of *Bacteroides* and *Akkermansia* associated with microbial Q-process pathways in UC. Forest plots quantify the multi-cohort poolings of SMD for microbial preQ_0_ biosynthesis biosynthesis pathway (highlighted in yellow) contributed by **(A)** *B. dorei*, (B) *B. plebeius,* **(C)** *A. muciniphila*, and Q-biosynthesis pathway (highlighted in blue) consistent down-regulation contributed by specific bacteria species **(D)** *B. dorei*, **(E)** *B. massiliensis*, **(F)** *A. muciniphila* in UC patients. For each individual study, squares denote the SMD, with their dimensions scaling according to the assigned study weight. The 95% CI are depicted by horizontal lines, while diamonds illustrate the combined effect sizes derived from random-effects and fixed-effects models. Inter-study heterogeneity is reflected by the I^2^ and p-values detailed at the base of each panel. Pooled P values of both models are shown in each panel. Regarding clinical interpretations, negative SMD values signify a reduction in UC samples, whereas positive values denote enrichment. Each panel includes the studies with detectable contributions, n = 1740 (830 UC and 910 healthy controls).

### Mixed-effects modelling confirms key taxonomic shifts in Q biosynthesis processes

To systematically characterize individual key microbial species in IBD, across the features analyzed in Figure 3, 4, S1, S2, after controlling for potential confounders including sex, age and study batch effects, the relative abundance of individual target bacteria species and pathways still showed significant differences between IBD patients (CD and UC) and healthy controls (**data not shown**). After adjustment, the direction of association was unchanged in all 29 models, and diagnosis remained significantly associated with abundance in 22 (11/13 in CD, 11/16 in UC). In CD, the depletion of *B. massiliensis (***Fig. 3B, E**) and the enrichment of *Escherichia coli* (**Fig. S1D**) were independent of age, but conditioned on male. The depletion of *Bacteroides plebeius* (**Fig. 3D, G**), *B._intestinihominis* (**Fig. S1E**), *B. dorei* (**Fig.S1B**) and *A._putredinis* (**Fig. S1F**) were associated with age and conditioned on male. In UC, there was changes of *B. dorei*, *B. massiliensis*, *B. xylanisolvens*, *B. intestinihominis*, *Alistipes* spp. and *A. muciniphila* in the Q biosynthesis pathway (**Fig. 4 UC**). Sex was a strong determinant of abundance, and a diagnosis × sex interaction was significant in 17 of 29 models, with the depletion of *Bacteroides* species (e.g., *B. massiliensis*, *B. plebeius*, *B. dorei*) being more pronounced in males. Study heterogeneity was significant in 19 of 29 models (**data not shown**).

### Female predominance of q-producing bacteria in CD and UC and age-dependent decline of q-producing bacteria in UC

The mixed-effects models also showed that sex and age were significant determinants of the abundance of many q-producing taxa (sex, 20 of 29 models; age, 18 of 29 models; **data not shown**), and that sex modified the diagnosis effect in 16 of 29 models, indicating that host physiological factors shape the q-producing community beyond disease status. We therefore examined the aggregate abundance of key q-producing bacteria (*Bacteroides* and *Alistipes*) according to sex and age. The combined abundance was significantly higher in females than in males in both CD and UC (**Fig. 5A**, **5B**), although the direction of species-level sex effects varied (**Data not shown**). Among 750 CD and 459 UC samples, host age at diagnosis was weakly but significantly negatively correlated with the combined abundance in UC, not in CD (Fig. 5C, 5D), consistent with the negative age coefficients for *A. putredinis* and *A. senegalensis* in UC, although age coefficients were positive for several Bacteroides species and for *A. indistinctus* and *A. shahii* (**data not shown**). These findings suggest that males and older individuals may harbour a less robust Q-biosynthetic bacterial community, which could contribute to demographic differences in IBD susceptibility or progression.

**Figure 5.**
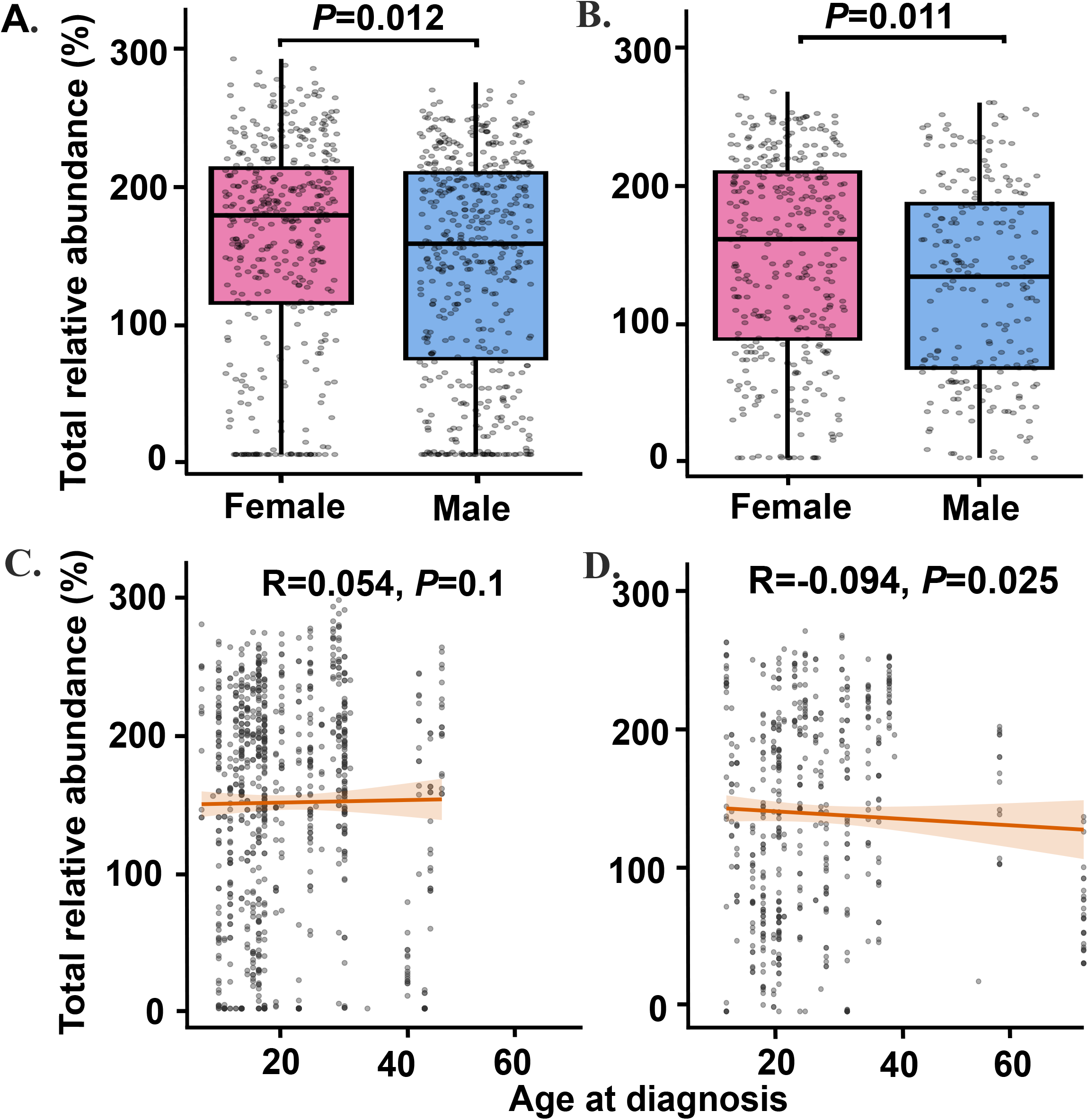
q-producing bacteria in UC decreased with age and are higher in female than male patients in both CD and UC. Sex differences in the abundance of q-producing bacteria in **(A)** CD and **(B)** UC patients. Box plots show the total relative abundance of major q-producing genera (*Bacteroides* and *Alistipes*) (sum of species-level relative abundances per sample) in female and male cohorts in CD and UC patients. Females showed significant higher total abundance compared to males. Data are presented as medians with interquartile ranges (IQR), each dot represents an individual sample. Age-associated shifts in q-producing bacteria in **(C)** CD and **(D)** UC patients. Spearman correlation revealed the association between the total relative abundance of q-producing bacteria and age at diagnosis in CD and UC patients. A weak but significant negative correlation suggests a gradual decline of the q-producing bacteria with elder age in UC patients. The orange line represents the linear regression fit with a 95% CI shaded in light orange. Wilcoxon rank sum test, n=1638 samples (750 CD, 459 UC, 429 healthy controls).

### Downregulated QTRT1 expression and convergence of related pathways

Beyond its established role in tRNA modification, the queuosine biosynthesis and regulatory network in *Bacteroides uniformis* appears to be integrated with metabolism based on the published study Kuziel et al. (Cell, 2024)^23^. Our re-analysis of the Q-pathway genes demonstrated that these genes contribute to fitness during the catabolism of phenolic glycosides such as arbutin (**Fig. 6A**). Specifically, TGT fitness requirement increased in *Bacteroides uniformis* ΔgshD (arbutin environment), and Q-tRNA-modification dependence increased for arbutin utilization. These findings suggest that the Q-related metabolic machinery in the gut microbiota may have evolved to facilitate the diversification of dietary small molecules, potentially linking micronutrient q-availability to the metabolic processing of inflammatory process. scRNA-seq data, visualized via Uniform Manifold Approximation and Projection (UMAP), identified a predominantly enriched QTRT1 cluster in IECs, with lower signal in other cell types (**Fig. 6B**). To determine the clinical relevance of QTRT1 in epithelial cells of IBD patients, we reanalyzed scRNA-seq datasets in NCBI-GEO. We found the gene expression level of QTRT1 in IECs was significantly downregulated in IBD patients compared with healthy controls (**Fig. 6C**). This significant downregulation suggests that the loss of QTRT1 may be a characteristic feature of the dysfunctional intestinal epithelium in IBD. To further elucidate the biological impact of QTRT1 downregulation in IECs, we performed Gene Ontology (GO) enrichment analysis on the differentially expressed genes associated with IBD epithelial dysfunction. Our analysis showed that genes upregulated in IBD IECs were mainly enriched in processes related to cell adhesion, intrinsic apoptotic signaling, and endoplasmic reticulum (ER) stress. In contrast, downregulated genes were enriched in protein synthesis and energy metabolism (**Fig. 6D**).

**Figure 6.**
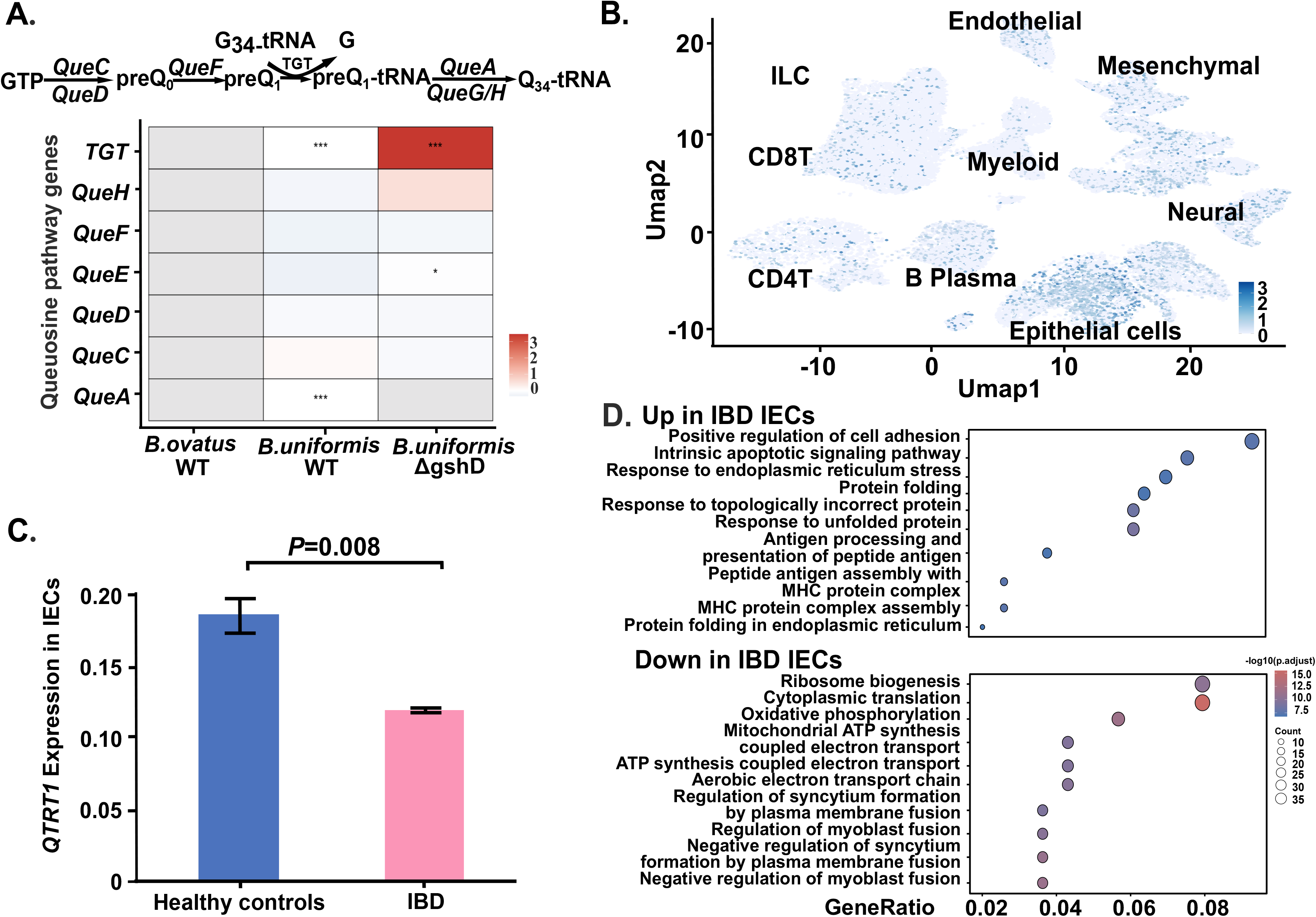
Downregulation of QTRT1 in host IECs and bacterial Q-related genes is associated with dysregulated immune and bacterial response pathways in IBD. **(A)** Data were curated from Kuziel et al. (Cell, 2024) and re-analyzed to evaluate the role of Q-related machinery in small molecule metabolism. The complete growth arrest in ΔgshD suggests a critical link between Q-associated genomic loci and phenolic glycoside catabolism. Data are shown as mean ± standard deviation (SD), Two-way ANOVA. **(B)** UMAP visualization of scRNA-seq data highlighting the clustering and distribution of QTRT1 gene in the IECs population. **(C)** QTRT1 expression was significantly downregulated in colonic epithelial cells from patients with IBD compared with healthy controls as determined by scRNA-seq. The data were retrieved from NCBI-GEO scRNA-seq database, which included 43,692 cells from three CD patients and 51,036 cells from four healthy controls. Data are shown as mean ± SD, Welch’s t-test. **(D)** GO biological process term enriched among genes up- and down-regulated in IBD compared with healthy control epithelial cells. The x-axis represents the Gene Ratio, while the dot size indicates the gene count and color gradient represents the adjusted *P*-value.

## Discussion

This study aims to advance our understanding of the biological roles of q-producing gut microbiota in healthy and IBD populations by applying the uniform bioinformatic pipeline to published datasets. We provide a comprehensive multi-omics characterization of the Q-axis in human IBD, specifically identifying a novel state of functional dysbiosis. Our meta-analysis of publicly available metagenomic datasets including 3146 samples reveal a systematic depletion of Q-biosynthetic potential in the IBD gut, driven by the loss of key q-producing taxa such as *Bacteroides* and *Alistipes*. By integrating these findings with scRNA-seq data, we establish that this microbial deficiency correlates with altered host QTRT1 expression, suggesting a profound breakdown in the trans-kingdom supply chain of this essential epitranscriptomic precursor. By integrating multi-omics data, our findings elucidate the functional microbial signatures driving CD and UC pathogenesis and provide a robust framework for identifying metabolic pathways amenable to therapeutic modulation.

To our knowledge, this is the first study to link gut microbial function with Q-modification of tRNA. Given that Q is indispensable for translational fidelity and mucosal proteostasis, disruption of this microbial Q-biosynthetic axis may be a fundamental driver of epithelial dysfunction in IBD. We assemble large sequencing datasets that was systematically curated, rigorously analyzed, and updated to current taxonomic standards, allowing us to characterize a functional dysbiosis in IBD marked by depletion of q-producing bacteria. IBD, comprising CD and UC, represents a complex breakdown of the homeostatic dialogue between host genetics, the gut microbiome, and dietary exposures, which imposes a profound biological and socioeconomic burden, evidenced by a significant reduction in life expectancy of five to eight years^17,18^. Our data have demonstrated the microbial Q-synthesis host tRNA modification axis as a critical mechanistic link in human IBD.

Whereas the reduction in microbial diversity and the role of specific pathogens in IBD have been established since the earliest culture-based studies in 1968, the field has largely remained focused on taxonomic associations^24–28^. Our study transcends these traditional descriptions by identifying a functional breakdown in the microbial-host Q supply chain for tRNA mofification. By integrating multi-omics data with single-cell resolution, we demonstrate that IBD pathogenesis is driven not just by the presence of harmful bacteria, but by the critical loss of q-producers. The functional importance of Bacteroides in this context is underscored by recent studies linking *B. vulgatus* and *B. fragilis* proteases to barrier disruption and intestinal inflammation^29,30^. Beyond proteolysis, Bacteroides act as essential metabolic hubs that liberate bioactive effectors to maintain homeostasis^23^. Consequently, the disrupted Q-biosynthesis created by dysbiosis represents a profound breakdown in this trans-kingdom supply chain, may drive epitranscriptomic instability in the intestinal epithelium.

Our findings in CD and UC have shown the abundance of q-producing bacteria is higher in females than in males. Previous research has demonstrated that while plasma q levels remain stable across different ages in healthy controls, they exhibit a significant sex-dependent variation, with higher levels observed in women than in men^31^. Circulating q, a gut-derived precursor essential for Q biosynthesis, is reported to decline with age in rodents and humans, accordingly, vitamin q supplementation extends lifespan across multiple dimensions^32^The implications of a disrupted Q-axis likely extend beyond the gastrointestinal tract. Emergent evidence suggests that Q-modification plays a role in systemic processes, including aging and neurodegeneration. Given that the gut microbiome is the primary source of systemic q, the functional dysbiosis observed here might serve as a causal link between intestinal inflammation and extra-intestinal manifestations, such as neuroinflammatory or metabolic disorders. We speculate that a targeted disruption in the gut microbiome’s queuosine biosynthetic capacity contributes to human IBD by impairing host tRNA modification and subsequent epithelial function. Investigating these causal pathways, particularly regarding sex-dimorphic microbiomes and their impact on autoimmunity, remains a high-priority area for future research. To address in methodologic inconsistency, our study leverages a unified bioinformatics approach across integrated datasets to elucidate the gut microbiota’s role in IBD. By rigorously updating taxonomy and focusing on the Q-biosynthesis pathway, we identify core microbial perturbations at disease onset. Notably, our analysis reveals a distinct sexual dimorphism in q-producing bacteria that mirrors systemic variations in healthy cohorts.

Our study represents the first large-scale investigation to systematically characterize the microbial-derived Q axis across a comprehensive IBD cohort. By applying a standardized bioinformatic pipeline, integrating HUMAnN3 for functional metagenomics and Seurat for single-cell transcriptomics, we have moved beyond simple taxonomic descriptions to define a novel functional signature of the disease. Harnessing the collective power of datasets encompassing over 3,000 samples from variable races and positions, our work provides the first high-resolution evidence that the disrupted Q is a hallmark of IBD. Beyond IBD, our framework highlights the value of harmonizing sequence data across international repositories and offers a transferable strategy that can be applied to other diseases to improve the efficiency of data analysis.

Despite the robust associations identified, our study is primarily based on computational meta-analyses. Vitamin q and its tRNA modification is lesser-recognized in human IBD and there are no public available data to measure the q level in patients with IBD. While the observation of microbial q-loss and host QTRT1 deficiency is striking, further experimental validation using human samples for q metabolites, germ-free mice or human intestinal organoids is required to definitively prove the causal impact of q-deficiency on epithelial ribosomal kinetics.

In conclusion, as illustrated in our Graphical Abstract, we have identified the loss of the microbial Q-axis as a hallmark of functional dysbiosis in human IBD. It advances our understanding of how the microbiome fine-tunes host in human health and IBD. Restoring this trans-kingdom epitranscriptomic link offers a promising strategy for achieving sustained mucosal homeostasis anti-chronic inflammation.

## Methods

### Publicly available CD and UC datasets collection

Google Scholar and PubMed searches were performed for microbiome studies in IBD including both CD and UC. Of 64 datasets assessed for eligibility including 51 eligible scRNA-seq datasets, 12 metagenomic datasets and one public metagenomic dataset, 25 were included in the bioinfomatic reanalysis including 9 metagenomic, 15 scRNA-seq datasets and one metagenomic dataset. 3146 samples in metagenomic datasets were included, comprising 1406 from CD, 830 from UC and 910 healthy controls. While 519 samples were screened from scRNA-seq datasets, including 139 CD samples, 172 UC samples and 208 healthy controls. Available metagenomics and scRNA sequence datasets were downloaded. Integrated data were run through a unified HUMAnN3 v3.9^22^, (coupled with MetaPhlAn 4 and MetaCyc v24.0 pathway definitions), species-stratified pathway abundances were expressed as copies per million, and Seurat v5.0 ^33^ bioinformatics pipeline.

### Meta-analysis of human gut microbiota metagenomic studies including bacteria species and functional pathways

Meta-analysis was using nine human gut microbiota metagenomic datasets of IBD patients compared with healthy controls^27,34–41^. The analysis of bacterial relative abundance variations between IBD patients and healthy controls were calculated at species level. For each pathway, the contribution of species was calculated per sample as its species-stratified pathway abundance divided by the total abundance of the same pathway in that sample. To evaluate the consistency of genomic abundance changes in specific bacterial functional pathways across different patient populations, we performed a meta-analysis for each identified bacteria taxa and pathways, the mean abundance, standard deviation, and sample size of these contributions were extracted for each study for both CD, UC and healthy controls using R v4.6.1 and Python v3.13. The meta-analysis was conducted using both fixed-effect model and random-effect model to account for potential heterogeneity across studies, such as differences in geography, patient recruitment, and sequencing depth. The Standardized Mean Difference (SMD) (Hedges’g) with its 95% confidence interval (CI) was employed as the effect size measure to quantify the difference in pathway abundance between groups. Between-study variance (τ²) was estimated by restricted maximum likelihood, and random-effects confidance intervals were Wald-type. Statistical heterogeneity among studies was assessed using the I^2^ statistic and the Cochran’s Q test. All statistical analyses and forest plot visualizations were performed using the meta package (metacont function) in R v4.6.1 software.

### Integrated analysis of human IBD scRNA-seq studies and metabolic studies

To investigate the clinical significance of QTRT1 in inflammatory bowel disease (IBD), we screened 15 publicly available IBD-related scRNA-seq data from the NCBI Gene Expression Omnibus database (https://www.ncbi.nlm.nih.gov/geo/). Among these datasets, GSE164985 was identified as a key resource containing colonic epithelial biopsy samples from 3 CD patients and 4 healthy controls, enabling evaluation of QTRT1 expression within disease-affected epithelial compartments^42^. Filtered 10x Genomics matrices of each sample were loaded and merged in the standardized pipeline Seurat v5.0. QTRT1 Unique Molecular Identifier (UMI) counts were averaged per sample and compared between IBD and healthy controls using Welch’s *t*-test. Differential expression between IBD and healthy controls was tested with Bonferroni-adjusted and GO biological process enrichment of up- and down-regulated genes was performed with clusterProfilter. We analyzed the synthetic Tn-seq data from Cell 2024 to identify genetic components co-operating with gshD for aryl glycoside metabolism in *B. uniformis*. The screen was performed on a ΔgshD background under arbutin selection. Significant fitness determinants were identified by filtering for genes with substantial depletion in the transposon library based on Log_2_FC.

### Statistical analyses

Each dataset was stratified by participant health status, categorized into healthy control, CD, and UC groups. Missing values were excluded prior to analysis. Metagenomic functional profiles were normalized by dividing species-specific abundances by the total pathway capacity to determine proportional contributions. Inter-group differences between healthy control, CD, and UC cohorts were evaluated using the Welch’s *t*-test for the species-level comparisons with mean difference ± SEM and Wilcoxon rank-sum test for sex comparison. To assess sex-dimorphic and age-associated shifts, Spearman’s rank correlation and multivariate linear regression were performed. All *P*-values were adjusted using the Benjamini-Hochberg FDR procedure. All statistical analyses were performed using R software v4.6.1.

### Mixed-effects modelling

To assess whether differences between IBD patients and healthy controls were independent of confounders, species-level contributions to each pathway were analyzed using linear mixed-effects models, lme4 and lmerTest packages in R v4.6.1^43^. Samples with complete data on diagnosis, sex, age and study were analyzed separately for CD versus healthy controls and UC versus healthy controls. Models were fitted by maximum likelihood with diagnosis (healthy controls as reference), sex and age as fixed effects and study as a random intercept (Abundance ∼ Diagnosis + Sex + Age + (1 | Study)). A second model additionally included a diagnosis × sex interaction. Backward stepwise elimination (lmerTest:step, random effects, α = 0.1, fixed effects, α = 0.05) was applied to the interaction model to obtain a reduced model. *P*-values for fixed effects were based on Satterthwaite’s approximation, and *P* < 0.05 was considered statistically significant.

### Conflict of interest

The authors declare no conflict of interest. The funders played no role in the study design, the collection, analyses, or interpretation of data, the writing of the manuscript, or the decision to publish the results.

## Data Availability

All data produced in the present study are available upon reasonable request to the authors

## Acknowledgements/Funds

We would like to acknowledge the Crohn’s & Colitis Foundation Senior Research Award (Grant No. 902766), the NIDDK/National Institutes of Health grant R01 DK134343, R01DK114126, VA Merit Award VA 1 I01 BX004824-06, and VA Collaborative Merit Award 1 I01BX006878-01A1 to Jun Sun. The study sponsors play no role in the study design, data collection, analysis, and interpretation of data. The contents do not represent the views of the United States Department of Veterans Affairs or the United States Government. Partial of our data was presented at the American Physiological Society (APS) 2026.

## Abbreviation list

ANOVA: analysis of variance
CD: Crohn’s disease
CI: confidence intervals
EMBL-EBI: European Molecular Biology Laboratory-European Bioinformatics Institute
ER: endoplasmic reticulum
GO: gene ontology
gsh: glycoside specialist hydrolase
GTP: Guanosine triphosphate
HUMAnN3: HMP unified metabolic analysis network 3
LMM: linear mixed-effects model
L_2_FC: log_2_ fold change
SD: standard deviation
MetaCyc: MetaCyc metabolic pathway database
MetaPhlAn 4: Metagenomic phylogenetic analysis 4
NCBI-GEO: National Center for Biotechnology Information-Gene Expression Omnibus
IBD: Inflammatory bowel disease
IBDMDB: The Inflammatory Bowel Disease Multi’omics Database
IECs: Intestinal epithelial cells
q: queuine
Q: queuosine
QTRT1: Queuine tRNA-ribosyltransferase 1
QTRT2: Queuine tRNA-ribosyltransferase 2
Q-tRNA: Queuosine-containing tRNA
SMD: standardized mean difference
scRNA-seq: single-cell RNA sequencing
TGT: tRNA-guanine transglycosylase
tRNA: transfer RNA
UC: Ulcerative colitis
UMAP: Uniform Manifold Approximation and Projection

---

**Figure S1. Divergent shifts in microbial Q-process pathways characterized by *Bacteroides*/*Alistipes* depression and concurrent *Escherichia coli* enrichment in CD.** Forest plots illustrate the SMD in genomic abundance for preQ_0_ biosynthesis pathway (highlighted in yellow) contributed by specific bacterial taxa (**A**) *B. intestinihominis*, (**B**) *B. dorei*, (**C**) *E. coli*, Q-biosynthesis pathway contributed by (**D**) *E. coli*, (**E**) *B. intestinihominis*, de novo guanosine ribonucleotide biosynthesis pathway (highlighted in blue) contributed by (**F**) *A. putredinis*, and de novo guanosine deoxyribonucleotide biosynthesis pathway (highlighted in pink) contributed (**G**) *A. putredinis* in CD patients. Squares represent the SMD for individual studies, with the size proportional to the study weight. Horizontal lines indicate the 95% CI. Diamonds represent the pooled effect size calculated via the random-effects and fixed-effects models. Heterogeneity is indicated by the I^2^ value and p-value provided at the bottom of each panel. Pooled *P* values of both models are shown in each panel. Negative SMD values indicate depletion in CD patients, while positive values indicate enrichment. Each panel includes the studies with detectable contributions, n=2316 samples (1406 CD, 910 healthy controls).

**Figure S2. Decrese of *Bacteroides* and *Alistipes* associated with microbial Q-process pathways in UC.** Forest plots quantify the multi-cohort poolings of Standardized Mean Difference (SMD) for microbial preQ_0_ biosynthesis biosynthesis pathway (highlighted in yellow) contributed by **(A)** *B. massiliensis*, (B) *B. eggerthii,* **(C)** *B. xylanisolvens*, **(D)** *B. intestinihominis*, Q-biosynthesis pathway (highlighted in blue) consistent down-regulation contributed by specific bacteria species **(E)** *B. xylanisolvens*, **(J)** *B. eggerthii*, and de novo guanosine ribonucleotide biosynthesis pathway (highlighted in pink) contributed by (**F**) *A. putredinis*, (**G**) *A. senegalensis*, (**H**) *A. indistinctus*, (**I**) *A. shahii* in UC patients. For each individual study, squares denote the SMD, with their dimensions scaling according to the assigned study weight. The 95% CI are depicted by horizontal lines, while diamonds illustrate the combined effect sizes derived from random-effects and fixed-effects models. Inter-study heterogeneity is reflected by the I^2^ and *P*-values detailed at the base of each panel. Pooled *P*-values of both models are shown in each panel. Regarding clinical interpretations, negative SMD values signify a reduction in UC samples, whereas positive values denote enrichment. Each panel includes the studies with detectable contributions, n = 1740 (830 UC and 910 healthy controls).

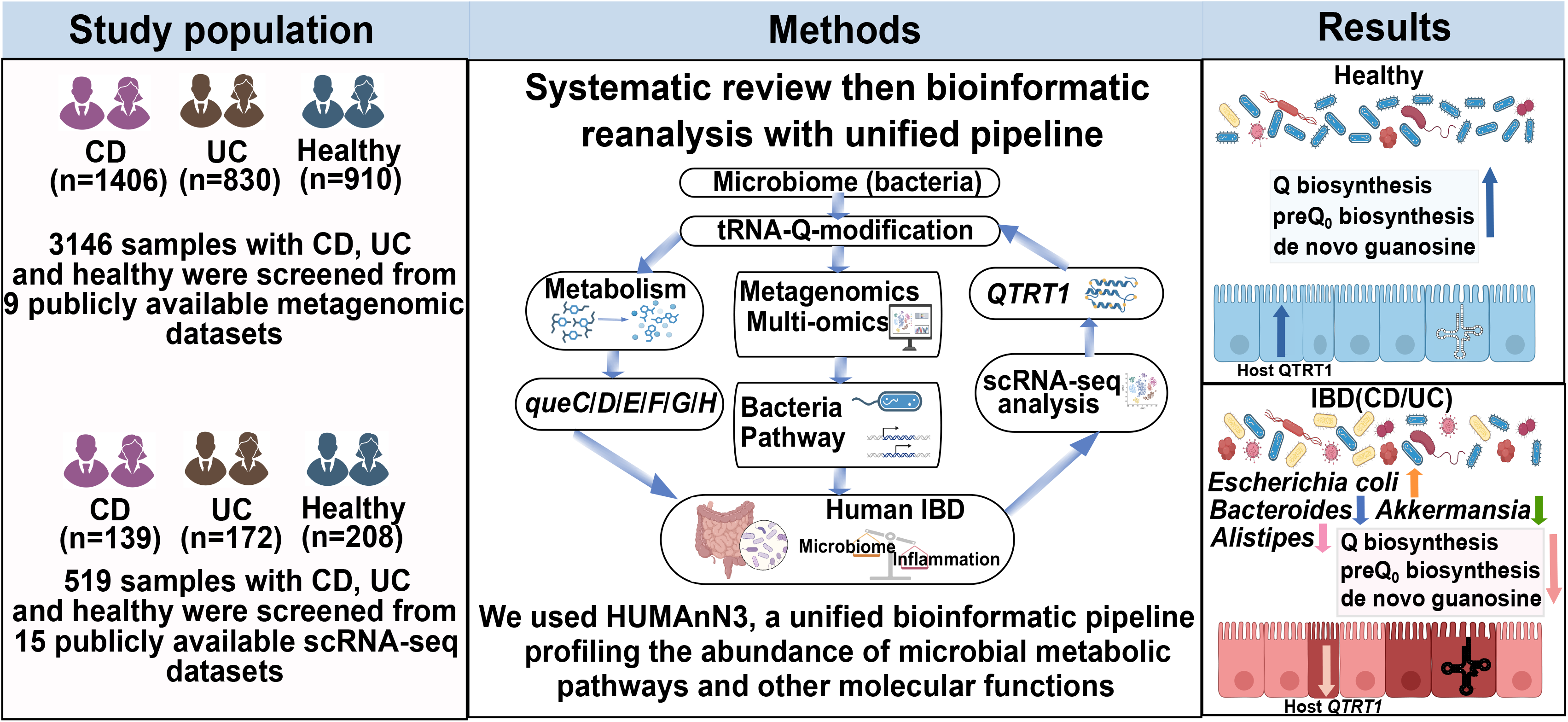

## Notes

### Competing Interest Statement

The authors have declared no competing interest.

## References

1. de Crécy-Lagard V, Hutinet G, Cediel-Becerra JD, Yuan Y, Zallot R, Chevrette MG, Ratnayake RMN, Jaroch M, Quaiyum S, Bruner S. Biosynthesis and function of 7-deazaguanine derivatives in bacteria and phages. Microbiology and Molecular Biology Reviews. 2024;88:e00199–00123. doi:

2. Fergus C, Barnes D, Alqasem MA, Kelly VP. The queuine micronutrient: charting a course from microbe to man. Nutrients. 2015;7:2897–2929. doi:

3. Sievers K, Welp L, Urlaub H, Ficner R. Structural and functional insights into human tRNA guanine transglycosylase. RNA biology. 2021;18:382–396. doi:

4. Sievers K, Neumann P, Sušac L, Da Vela S, Graewert M, Trowitzsch S, Svergun D, Tampé R, Ficner R. Structural and functional insights into tRNA recognition by human tRNA guanine transglycosylase. Structure. 2024;32:316–327. e315. doi:

5. Suzuki T. The expanding world of tRNA modifications and their disease relevance. Nature Reviews Molecular Cell Biology. 2021;22:375–392. doi:

6. Matsumura Y, Wei F-Y, Sakai J. Epitranscriptomics in metabolic disease. Nature Metabolism. 2023;5:370–384. doi:

7. Bento-Abreu A, Jager G, Swinnen B, Rue L, Hendrickx S, Jones A, Staats KA, Taes I, Eykens C, Nonneman A. Elongator subunit 3 (ELP3) modifies ALS through tRNA modification. Human molecular genetics. 2018;27:1276–1289. doi:

8. Zhang Y, Xu W, Peng C, Ren S, Hidig SM, Zhang C. Exploring the role of m7G modification in Cancer: Mechanisms, regulatory proteins, and biomarker potential. Cellular Signalling. 2024;121:111288. doi:

9. Dedon PC, Begley TJ. Dysfunctional tRNA reprogramming and codon-biased translation in cancer. Trends in molecular medicine. 2022;28:964–978. doi:

10. Añazco-Guenkova AM, Miguel-López B, Monteagudo-García Ó, García-Vílchez R, Blanco S. The impact of tRNA modifications on translation in cancer: identifying novel therapeutic avenues. NAR cancer. 2024;6:zcae012. doi:

11. Wang Y, Tao E-W, Tan J, Gao Q-Y, Chen Y-X, Fang J-Y. tRNA modifications: insights into their role in human cancers. Trends in Cell Biology. 2023;33:1035–1048. doi:

12. Santos MCFd, Anderson CP, Neschen S, Zumbrennen-Bullough KB, Romney SJ, Kahle-Stephan M, Rathkolb B, Gailus-Durner V, Fuchs H, Wolf E. Irp2 regulates insulin production through iron-mediated Cdkal1-catalyzed tRNA modification. Nature communications. 2020;11:296. doi:

13. Wei F-Y, Suzuki T, Watanabe S, Kimura S, Kaitsuka T, Fujimura A, Matsui H, Atta M, Michiue H, Fontecave M. Deficit of tRNA Lys modification by Cdkal1 causes the development of type 2 diabetes in mice. The Journal of clinical investigation. 2011;121. doi:

14. Asano K, Suzuki T, Saito A, Wei F-Y, Ikeuchi Y, Numata T, Tanaka R, Yamane Y, Yamamoto T, Goto T. Metabolic and chemical regulation of tRNA modification associated with taurine deficiency and human disease. Nucleic Acids Res. 2018;46:1565–1583. doi:

15. Chen D, Li F, Yang Q, Tian M, Zhang Z, Zhang Q, Chen Y, Guan M-X. The defective expression of gtpbp3 related to tRNA modification alters the mitochondrial function and development of zebrafish. The international journal of biochemistry & cell biology. 2016;77:1–9. doi:

16. Zhang J, Zhang Y, McGrenaghan CJ, Kelly VP, Xia Y, Sun J. Disruption to tRNA Modification by Queuine Contributes to Inflammatory Bowel Disease. Cell Mol Gastroenterol Hepatol. 2023;15:1371–1389. doi: 10.1016/j.jcmgh.2023.02.006

17. Lewis JD, Parlett LE, Funk MLJ, Brensinger C, Pate V, Wu Q, Dawwas GK, Weiss A, Constant BD, McCauley M. Incidence, prevalence, and racial and ethnic distribution of inflammatory bowel disease in the United States. Gastroenterology. 2023;165:1197–1205. e1192. doi:

18. Kuenzig ME, Manuel DG, Donelle J, Benchimol EI. Life expectancy and health-adjusted life expectancy in people with inflammatory bowel disease. Cmaj. 2020;192:E1394–E1402. doi:

19. Marks T, Farkas W. Effects of a diet deficient in tyrosine and queuine on germfree mice. Biochemical and biophysical research communications. 1997;230:233–237. doi:

20. Ehrenhofer-Murray AE. Queuine: a bacterial nucleobase shaping translation in eukaryotes. Journal of Molecular Biology. 2025;437:168985. doi:

21. Díaz-Rullo J, González-Pastor JE. tRNA queuosine modification is involved in biofilm formation and virulence in bacteria. Nucleic Acids Res. 2023;51:9821–9837. doi:

22. Beghini F, McIver LJ, Blanco-Míguez A, Dubois L, Asnicar F, Maharjan S, Mailyan A, Manghi P, Scholz M, Thomas AM. Integrating taxonomic, functional, and strain-level profiling of diverse microbial communities with bioBakery 3. elife. 2021;10:e65088. doi:

23. Kuziel GA, Lozano GL, Simian C, Li L, Manion J, Stephen-Victor E, Chatila T, Dong M, Weng J-K, Rakoff-Nahoum S. Functional diversification of dietary plant small molecules by the gut microbiome. Cell. 2025;188:1967–1983. e1922. doi:

24. Hawkins HP. An address on the natural history of ulcerative colitis and its bearing on treatment. British Medical Journal. 1909;1:765. doi:

25. Gevers D, Kugathasan S, Denson LA, Vázquez-Baeza Y, Van Treuren W, Ren B, Schwager E, Knights D, Song SJ, Yassour M. The treatment-naive microbiome in new-onset Crohn’s disease. Cell host & microbe. 2014;15:382–392. doi:

26. Pérez-Brocal V, García-López R, Nos P, Beltrán B, Moret I, Moya A. Metagenomic analysis of Crohn’s disease patients identifies changes in the virome and microbiome related to disease status and therapy, and detects potential interactions and biomarkers. Inflammatory Bowel Diseases. 2015;21:2515–2532. doi:

27. Lloyd-Price J, Arze C, Ananthakrishnan AN, Schirmer M, Avila-Pacheco J, Poon TW, Andrews E, Ajami NJ, Bonham KS, Brislawn CJ. Multi-omics of the gut microbial ecosystem in inflammatory bowel diseases. Nature. 2019;569:655–662. doi:

28. Rojas-Feria M, Romero-García T, Caballero-Rico JÁF, Ramírez HP, Avilés-Recio M, Castro-Fernandez M, Porcuna NC, Romero-Gόmez M, García F, Grande L. Modulation of faecal metagenome in Crohn’s disease: Role of microRNAs as biomarkers. World Journal of Gastroenterology. 2018;24:5223. doi:

29. Mills RH, Dulai PS, Vázquez-Baeza Y, Sauceda C, Daniel N, Gerner RR, Batachari LE, Malfavon M, Zhu Q, Weldon K. Multi-omics analyses of the ulcerative colitis gut microbiome link Bacteroides vulgatus proteases with disease severity. Nature microbiology. 2022;7:262–276. doi:

30. Lakemeyer M, Latorre R, Blazkova K, Wood HM, Jensen DD, Shakil N, Thomas SC, Saxena D, Mulpuri Y, Poolman D. A Bacteroides fragilis protease activates host PAR2 to induce intestinal pain and inflammation. Cell Host & Microbe. 2025;33:1686–1702. e1611. doi:

31. Richard P, Kozlowski L, Guillorit H, Garnier P, McKnight NC, Danchin A, Manière X. Queuine, a bacterial-derived hypermodified nucleobase, shows protection in in vitro models of neurodegeneration. PloS one. 2021;16:e0253216. doi:

32. Gong R-Z, Yan T-M, Pan Y, Cao K-Y, Cheng Y-T, Mo L-Y, Jiang Z-H. Evolutionarily Conserved Decline of tRNA Mannosyl-Queuosine Links Translational Regulation to Aging and Is Reversed by Queuine. bioRxiv. 2026:2026.2003. 2022.713446. doi:

33. Rich JM, Moses L, Einarsson PH, Jackson K, Luebbert L, Booeshaghi AS, Antonsson S, Sullivan DK, Bray N, Melsted P. The impact of package selection and versioning on single-cell RNA-seq analysis. Cell Systems. 2026;17. doi:

34. Norman JM, Handley SA, Baldridge MT, Droit L, Liu CY, Keller BC, Kambal A, Monaco CL, Zhao G, Fleshner P. Disease-specific alterations in the enteric virome in inflammatory bowel disease. Cell. 2015;160:447–460. doi:

35. Mancabelli L, Milani C, Lugli GA, Turroni F, Cocconi D, Van Sinderen D, Ventura M. Identification of universal gut microbial biomarkers of common human intestinal diseases by meta-analysis. FEMS microbiology ecology. 2017;93:fix153. doi:

36. Hall AB, Yassour M, Sauk J, Garner A, Jiang X, Arthur T, Lagoudas GK, Vatanen T, Fornelos N, Wilson R. A novel Ruminococcus gnavus clade enriched in inflammatory bowel disease patients. Genome medicine. 2017;9:103. doi:

37. Sankarasubramanian J, Ahmad R, Avuthu N, Singh AB, Guda C. Gut microbiota and metabolic specificity in ulcerative colitis and Crohn’s disease. Frontiers in medicine. 2020;7:606298. doi:

38. Pascal V, Pozuelo M, Borruel N, Casellas F, Campos D, Santiago A, Martinez X, Varela E, Sarrabayrouse G, Machiels K. A microbial signature for Crohn’s disease. Gut. 2017;66:813–822. doi:

39. Forbes JD, Chen C-y, Knox NC, Marrie R-A, El-Gabalawy H, de Kievit T, Alfa M, Bernstein CN, Van Domselaar G. A comparative study of the gut microbiota in immune-mediated inflammatory diseases—does a common dysbiosis exist? Microbiome. 2018;6:221. doi:

40. Franzosa EA, Sirota-Madi A, Avila-Pacheco J, Fornelos N, Haiser HJ, Reinker S, Vatanen T, Hall AB, Mallick H, McIver LJ. Gut microbiome structure and metabolic activity in inflammatory bowel disease. Nature microbiology. 2019;4:293–305. doi:

41. Schirmer M, Franzosa EA, Lloyd-Price J, McIver LJ, Schwager R, Poon TW, Ananthakrishnan AN, Andrews E, Barron G, Lake K. Dynamics of metatranscription in the inflammatory bowel disease gut microbiome. Nature microbiology. 2018;3:337–346. doi:

42. Kinchen J, Chen HH, Parikh K, Antanaviciute A, Jagielowicz M, Fawkner-Corbett D, Ashley N, Cubitt L, Mellado-Gomez E, Attar M, et al. Structural Remodeling of the Human Colonic Mesenchyme in Inflammatory Bowel Disease. Cell. 2018;175:372–386 e317. doi: 10.1016/j.cell.2018.08.067

43. Xia Y, Sun J. Bioinformatic and statistical analysis of microbiome data. In: From Raw Sequences to Advanced Modeling with QIIME 2 and R. Springer; 2023.

