## Supplemental data for "Disrupted microbial queuosine biosynthesis contributes to dysbiosis in human inflammatory bowel disease"

# CD

### preQ$_0$ biosynthesis pathway

#### A. *Bacteroides intestinihominis*

| Study | CD Mean | CD SD | CD Total | Healthy Mean | Healthy SD | Healthy Total |
|---|---|---|---|---|---|---|
| Imhann F et al., 2016 | 0.0032 | 0.0065 | 18 | 0.0051 | 0.0084 | 21 |
| Lewis JD et al., 2015 | 0.0061 | 0.0202 | 246 | 0.0130 | 0.0167 | 21 |
| Gever D et al., 2014 | 0.0051 | 0.0133 | 70 | 0.0086 | 0.0228 | 14 |
| Morgan XC., 2012 | 0.0011 | 0.0044 | 97 | 0.0069 | 0.0077 | 20 |
| Lloyd-Price J et al., 2019 | 0.0052 | 0.0162 | 762 | 0.0087 | 0.0158 | 449 |
| Li J et al., 2014 | 0.0097 | 0.0137 | 76 | 0.0070 | 0.0167 | 10 |
| He Q et al., 2017 | 0.0003 | 0.0023 | 49 | 0.0017 | 0.0043 | 53 |
| Total (fixed effect, 95% CI) | | | 1318 | | | 588 |
| Total (random effect, 95% CI) | | | | | | |

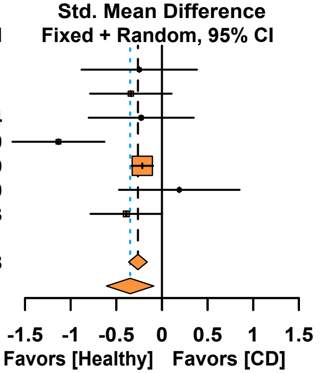


Heterogeneity: $Tau^2$ = 0.0677; $Chi^2$ = 14.48, df = 6 ($P$ = 0.0248); $I^2$ = 58.6%
Test for overall effect (fixed effect):Z= -5.06 ($P$< 0.0001)
Test for overall effect (random effects):Z= -2.63 ($P$= 0.0086)

#### B. *Bacteroides dorei*

| Study | CD Mean | CD SD | CD Total | Healthy Mean | Healthy SD | Healthy Total |
|---|---|---|---|---|---|---|
| Imhann F et al., 2016 | 0.0158 | 0.0341 | 18 | 0.0054 | 0.0111 | 21 |
| Lewis JD et al., 2015 | 0.0386 | 0.1099 | 246 | 0.0873 | 0.1378 | 21 |
| Gever D et al., 2014 | 0.0208 | 0.0552 | 70 | 0.0101 | 0.0248 | 14 |
| Morgan XC., 2012 | 0.0420 | 0.0703 | 97 | 0.0791 | 0.0920 | 20 |
| Lloyd-Price J et al., 2019 | 0.0435 | 0.1182 | 762 | 0.0583 | 0.1487 | 449 |
| Li J et al., 2014 | 0.0448 | 0.0776 | 76 | 0.0597 | 0.0667 | 10 |
| Hall AB et al., 2017 | 0.0503 | 0.0643 | 88 | 0.0622 | 0.0685 | 74 |
| He Q et al., 2017 | 0.0258 | 0.0664 | 49 | 0.0199 | 0.0479 | 53 |
| Total (fixed effect, 95% CI) | | | 1406 | | | 662 |
| Total (random effect, 95% CI) | | | | | | |

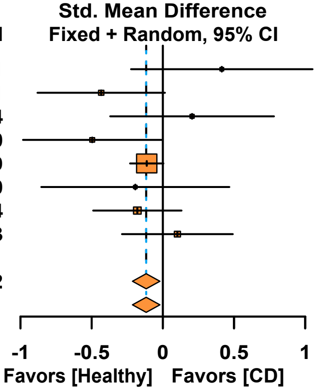


Heterogeneity: $Tau^2$ < 0.0001; $Chi^2$ = 9.57, df = 7 ($P$ = 0.2142); $I^2$ = 26.9%
Test for overall effect (fixed effect):Z= -2.38 ($P$= 0.0175)
Test for overall effect (random effects):Z= -2.37 ($P$= 0.0176)

#### C. *Escherichia coli*

| Study | CD Mean | CD SD | CD Total | Healthy Mean | Healthy SD | Healthy Total |
|---|---|---|---|---|---|---|
| Imhann F et al., 2016 | 0.0112 | 0.0228 | 18 | 0.0061 | 0.0188 | 21 |
| Lewis JD et al., 2015 | 0.0599 | 0.1386 | 246 | 0.0004 | 0.0008 | 21 |
| Gever D et al., 2014 | 0.0393 | 0.0984 | 70 | 0.0390 | 0.0644 | 14 |
| Morgan XC., 2012 | 0.0166 | 0.0596 | 97 | 0.0006 | 0.0027 | 20 |
| Lloyd-Price J et al., 2019 | 0.0217 | 0.0873 | 762 | 0.0074 | 0.0350 | 449 |
| He Q et al., 2017 | 0.1590 | 0.2383 | 49 | 0.0328 | 0.0690 | 53 |
| Total (fixed effect, 95% CI) | | | 1242 | | | 578 |
| Total (random effect, 95% CI) | | | | | | |

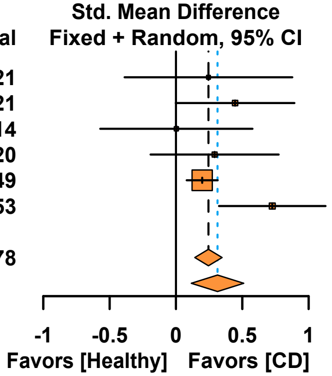


Heterogeneity: $Tau^2$ = 0.0217; $Chi^2$ = 7.63, df = 5 ($P$ = 0.1775); $I^2$ = 34.5%
Test for overall effect (fixed effect):Z= 4.66 ($P$< 0.0001)
Test for overall effect (random effects):Z= 3.10 ($P$= 0.0019)

### Q biosynthesis pathway

#### D. *Escherichia coli*

| Study | CD Mean | CD SD | CD Total | Healthy Mean | Healthy SD | Healthy Total |
|---|---|---|---|---|---|---|
| Imhann F et al., 2016 | 0.0117 | 0.0203 | 18 | 0.0049 | 0.0153 | 21 |
| Lewis JD et al., 2015 | 0.0498 | 0.1229 | 246 | 0.0002 | 0.0004 | 21 |
| Gever D et al., 2014 | 0.0386 | 0.0948 | 70 | 0.0336 | 0.0560 | 14 |
| Morgan XC., 2012 | 0.0119 | 0.0398 | 97 | 0.0007 | 0.0030 | 20 |
| Lloyd-Price J et al., 2019 | 0.0237 | 0.0899 | 762 | 0.0064 | 0.0277 | 449 |
| He Q et al., 2017 | 0.1440 | 0.2040 | 49 | 0.0240 | 0.0511 | 53 |
| Total (common effect, 95% CI) | | | 1242 | | | 578 |
| Total (random effect, 95% CI) | | | | | | |

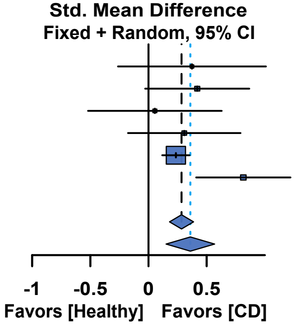


Heterogeneity: $Tau^2$ = 0.0266; $Chi^2$ = 8.32, df = 5 ($P$ = 0.1394); $I^2$ = 39.9%
Test for overall effect (fixed effect):Z= 5.40 ($P$< 0.0001)
Test for overall effect (random effects):Z= 3.38 ($P$= 0.0007)

#### E. *Bacteroides intestinihominis*

| Study | CD Mean | CD SD | CD Total | Healthy Mean | Healthy SD | Healthy Total |
|---|---|---|---|---|---|---|
| Imhann F et al., 2016 | 0.0025 | 0.0053 | 18 | 0.0051 | 0.0083 | 21 |
| Lewis JD et al., 2015 | 0.0061 | 0.0200 | 246 | 0.0126 | 0.0169 | 21 |
| Gever D et al., 2014 | 0.0044 | 0.0112 | 70 | 0.0084 | 0.0217 | 14 |
| Morgan XC., 2012 | 0.0011 | 0.0046 | 97 | 0.0067 | 0.0066 | 20 |
| Lloyd-Price J et al., 2019 | 0.0048 | 0.0151 | 762 | 0.0085 | 0.0170 | 449 |
| Li J et al., 2014 | 0.0073 | 0.0097 | 76 | 0.0036 | 0.0076 | 10 |
| Hall AB et al., 2017 | 0.0003 | 0.0002 | 49 | 0.0011 | 0.0026 | 53 |
| Total (fixed effect, 95% CI) | | | 1318 | | | 588 |
| Total (random effect, 95% CI) | | | | | | |

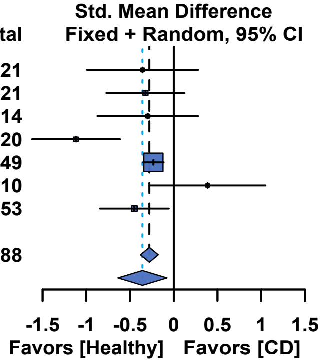


Heterogeneity: $Tau^2$ = 0.0869; $Chi^2$ = 16.01, df = 6 ($P$ = 0.0137); $I^2$ = 62.5%
Test for overall effect (fixed effect):Z= -5.36 ($P$< 0.0001)
Test for overall effect (random effects):Z= -2.49 ($P$= 0.0127)

### de novo guanosine ribonucleotide biosynthesis pathway

#### F. *Alistipes putredinis*

| Study | CD Mean | CD SD | CD Total | Healthy Mean | Healthy SD | Healthy Total |
|---|---|---|---|---|---|---|
| NL-IBD | 0.0033 | 0.0075 | 18 | 0.0085 | 0.0099 | 21 |
| Lewis JD et al., 2015 | 0.0287 | 0.0531 | 246 | 0.0336 | 0.0369 | 21 |
| Gever D et al., 2014 | 0.0084 | 0.0156 | 70 | 0.0084 | 0.0187 | 14 |
| Morgan XC., 2012 | 0.0137 | 0.0243 | 97 | 0.0322 | 0.0153 | 20 |
| Lloyd-Price J et al., 2019 | 0.0086 | 0.0142 | 762 | 0.0130 | 0.0141 | 449 |
| He Q et al., 2017 | 0.0035 | 0.0117 | 49 | 0.0050 | 0.0074 | 53 |
| Total (fixed effect, 95% CI) | | | 1242 | | | 578 |
| Total (random effect, 95% CI) | | | | | | |

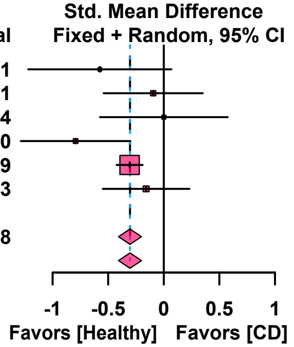


Heterogeneity: $Tau^2$ < 0.0001; $Chi^2$ = 6.94, df = 5 ($P$ = 0.2255); $I^2$ = 27.9%
Test for overall effect (fixed effect):Z= -5.77 ($P$< 0.0001)
Test for overall effect (random effects):Z= -5.77 ($P$< 0.0001)

### de novo guanosine deoxyribonucleotide biosynthesis pathway

#### G. *Escherichia coli*

| Study | CD Mean | CD SD | CD Total | Healthy Mean | Healthy SD | Healthy Total |
|---|---|---|---|---|---|---|
| Imhann F et al., 2016 | 0.0058 | 0.0099 | 18 | 0.0074 | 0.0303 | 21 |
| Lewis JD et al., 2015 | 0.0599 | 0.1386 | 246 | 0.0004 | 0.0008 | 21 |
| Gever D et al., 2014 | 0.0382 | 0.1005 | 70 | 0.0040 | 0.0070 | 14 |
| Morgan XC., 2012 | 0.0091 | 0.0482 | 97 | 0.0020 | 0.0064 | 20 |
| Lloyd-Price J et al., 2019 | 0.0355 | 0.1141 | 762 | 0.0135 | 0.0540 | 449 |
| He Q et al., 2017 | 0.2090 | 0.2569 | 49 | 0.0635 | 0.1132 | 53 |
| Total (fixed effect, 95% CI) | | | 1242 | | | 578 |
| Total (random effect, 95% CI) | | | | | | |

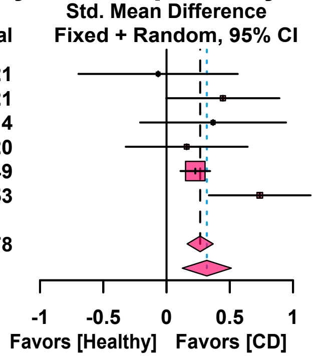


Heterogeneity: $Tau^2$ = 0.0201; $Chi^2$ = 7.70, df = 5 ($P$ = 0.1736); $I^2$ = 35.1%
Test for overall effect (fixed effect):Z= 5.06 ($P$< 0.0001)
Test for overall effect (random effects):Z= 3.22 ($P$= 0.0013)
