## Supplemental data for "Disrupted microbial queuosine biosynthesis contributes to dysbiosis in human inflammatory bowel disease"

# UC

### preQ$_0$ biosynthesis pathway

#### A. *Bacteroides massiliensis*

| Study | UC Mean | UC SD | UC Total | Healthy Mean | Healthy SD | Healthy Total |
|---|---|---|---|---|---|---|
| Imhann F et al., 2016 | 0.0035 | 0.0140 | 23 | 0.0040 | 0.0067 | 21 |
| Morgan XC., 2012 | 0.0141 | 0.0306 | 48 | 0.0164 | 0.0241 | 20 |
| Lloyd-Price J et al., 2019 | 0.0162 | 0.0382 | 471 | 0.0353 | 0.0793 | 449 |
| Hall AB et al., 2017 | 0.0067 | 0.0217 | 97 | 0.0057 | 0.0161 | 74 |
| Total (fixed effect, 95% CI) | | | 639 | | | 564 |
| Total (random effect, 95% CI) | | | | | | |

Heterogeneity: $Tau^2$ = 0.0247; $Chi^2$ = 5.50, df = 3 ($P$ = 0.1385); $I^2$ = 45.5%
Test for overall effect (fixed effect):Z= -4.06 ($P$< 0.0001)
Test for overall effect (random effects):Z= -1.24 ($P$= 0.2134)

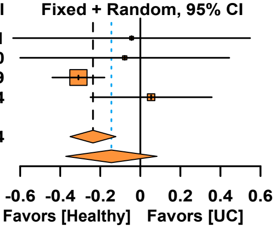


#### B. *Bacteroides eggerthii*

| Study | UC Mean | UC SD | UC Total | Healthy Mean | Healthy SD | Healthy Total |
|---|---|---|---|---|---|---|
| Imhann F et al., 2016 | 0.0036 | 0.0170 | 23 | 0.0021 | 0.0045 | 21 |
| Morgan XC., 2012 | 1.0e-9 | 1.0e-9 | 48 | 0.0221 | 0.0361 | 20 |
| Lloyd-Price J et al., 2019 | 0.0125 | 0.0457 | 471 | 0.0166 | 0.0513 | 449 |
| Hall AB et al., 2017 | 0.0001 | 0.0001 | 97 | 0.0061 | 0.0214 | 74 |
| Total (fixed effect, 95% CI) | | | 639 | | | 564 |
| Total (random effect, 95% CI) | | | | | | |

Heterogeneity: $Tau^2$ = 0.1959; $Chi^2$ = 16.70, df = 3 ($P$ = 0.0008); $I^2$ = 82.0%
Test for overall effect (fixed effect):Z= -2.92 ($P$= 0.0035)
Test for overall effect (random effects):Z= -1.49 ($P$= 0.1364)

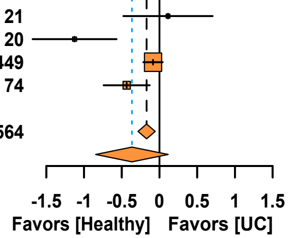


#### C. *Bacteroides xylanisolvens*

| Study | UC Mean | UC SD | UC Total | Healthy Mean | Healthy SD | Healthy Total |
|---|---|---|---|---|---|---|
| Imhann F et al., 2016 | 0.0020 | 0.0048 | 23 | 0.0010 | 0.0024 | 21 |
| Morgan XC., 2012 | 0.0022 | 0.0054 | 48 | 0.0022 | 0.0031 | 20 |
| Lloyd-Price J et al., 2019 | 0.0059 | 0.0196 | 471 | 0.0148 | 0.0250 | 449 |
| Nielsen HB et al., 2014 | 0.0015 | 0.0042 | 127 | 0.0044 | 0.0111 | 248 |
| Hall AB et al., 2017 | 0.0124 | 0.0420 | 97 | 0.0034 | 0.0093 | 74 |
| Total (fixed effect, 95% CI) | | | 766 | | | 812 |
| Total (random effect, 95% CI) | | | | | | |

Heterogeneity: $Tau^2$ = 0.0819; $Chi^2$ = 20.29, df = 4 ($P$ = 0.0004); $I^2$ = 80.3%
Test for overall effect (fixed effect):Z= -5.16 ($P$< 0.0001)
Test for overall effect (random effects):Z= -0.55 ($P$= 0.5809)

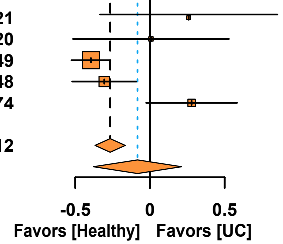


#### D. *Bacteroides intestinihominis*

| Study | UC Mean | UC SD | UC Total | Healthy Mean | Healthy SD | Healthy Total |
|---|---|---|---|---|---|---|
| Imhann F et al., 2016 | 0.0081 | 0.0133 | 23 | 0.0051 | 0.0084 | 21 |
| Morgan XC., 2012 | 0.0036 | 0.0105 | 48 | 0.0069 | 0.0077 | 20 |
| Lloyd-Price J et al., 2019 | 0.0034 | 0.0124 | 471 | 0.0087 | 0.0158 | 449 |
| Li J et al., 2014 | 0.0112 | 0.0128 | 64 | 0.0070 | 0.0167 | 10 |
| Total (fixed effect, 95% CI) | | | 606 | | | 500 |
| Total (random effect, 95% CI) | | | | | | |

Heterogeneity: $Tau^2$ = 0.0771; $Chi^2$ = 7.67, df = 3 ($P$ = 0.0535); $I^2$ = 60.9%
Test for overall effect (fixed effect):Z= -5.15 ($P$< 0.0001)
Test for overall effect (random effects):Z= -0.67 ($P$= 0.5050)

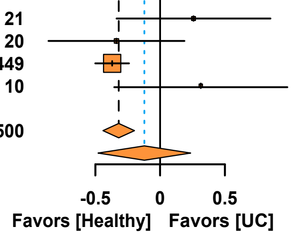


### Q biosynthesis pathway

#### E. *Bacteroides xylanisolvens*

| Study | UC Mean | UC SD | UC Total | Healthy Mean | Healthy SD | Healthy Total |
|---|---|---|---|---|---|---|
| Imhann F et al., 2016 | 0.0033 | 0.0079 | 23 | 0.0010 | 0.0019 | 21 |
| Morgan XC., 2012 | 0.0021 | 0.0049 | 48 | 0.0016 | 0.0025 | 20 |
| Lloyd-Price J et al., 2019 | 0.0079 | 0.0246 | 471 | 0.0140 | 0.0202 | 449 |
| Li J et al., 2014 | 0.0019 | 0.0067 | 127 | 0.0056 | 0.0116 | 248 |
| Hall AB et al., 2017 | 0.0090 | 0.0253 | 97 | 0.0049 | 0.0135 | 74 |
| Total (fixed effect, 95% CI) | | | 766 | | | 812 |
| Total (random effect, 95% CI) | | | | | | |

Heterogeneity: $Tau^2$ = 0.0665; $Chi^2$ = 14.69, df = 4 ($P$ = 0.0054); $I^2$ = 72.8%
Test for overall effect (fixed effect):Z= -4.02 ($P$< 0.0001)
Test for overall effect (random effects):Z= -0.43 ($P$= 0.6697)

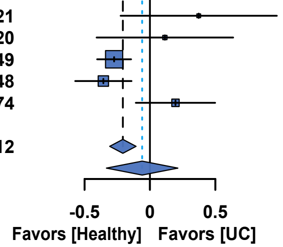


### de novo guanosine ribonucleotide biosynthesis pathway

#### F. *Alistipes putredinis*

| Study | UC Mean | UC SD | UC Total | Healthy Mean | Healthy SD | Healthy Total |
|---|---|---|---|---|---|---|
| Imhann F et al., 2016 | 0.0058 | 0.0085 | 23 | 0.0085 | 0.0099 | 21 |
| Morgan XC., 2012 | 0.0063 | 0.0164 | 48 | 0.0322 | 0.0153 | 20 |
| Lloyd-Price J et al., 2019 | 0.0063 | 0.0164 | 471 | 0.0130 | 0.0141 | 449 |
| Total (fixed effect, 95% CI) | | | 542 | | | 490 |
| Total (random effect, 95% CI) | | | | | | |

Heterogeneity: $Tau^2$ = 0.4082; $Chi^2$ = 14.45, df = 2 ($P$ = 0.0007); $I^2$ = 86.2%
Test for overall effect (fixed effect):Z= -7.52 ($P$< 0.0001)
Test for overall effect (random effects):Z= -1.90 ($P$= 0.0580)

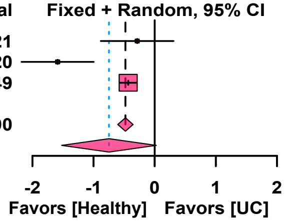


#### G. *Alistipes senegalensis*

| Study | UC Mean | UC SD | UC Total | Healthy Mean | Healthy SD | Healthy Total |
|---|---|---|---|---|---|---|
| Imhann F et al., 2016 | 0.0001 | 0.0005 | 23 | 0.0001 | 0.0005 | 21 |
| Morgan XC., 2012 | 0.0002 | 0.0006 | 48 | 0.0012 | 0.0023 | 20 |
| Total (fixed effect, 95% CI) | | | 71 | | | 41 |
| Total (random effect, 95% CI) | | | | | | |

Heterogeneity: $Tau^2$ = 0.1590; $Chi^2$ = 2.91, df = 1 ($P$ = 0.0879); $I^2$ = 65.7%
Test for overall effect (fixed effect):Z= -2.13 ($P$= 0.0335)
Test for overall effect (random effects):Z= -1.18 ($P$= 0.2390)

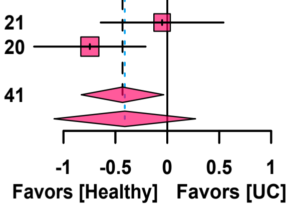


#### H. *Alistipes indistinctus*

| Study | UC Mean | UC SD | UC Total | Healthy Mean | Healthy SD | Healthy Total |
|---|---|---|---|---|---|---|
| Imhann F et al., 2016 | 0.0001 | 0.0002 | 23 | 0.0002 | 0.0005 | 21 |
| Morgan XC., 2012 | 0.0001 | 0.0003 | 48 | 0.0005 | 0.0007 | 20 |
| Lloyd-Price J et al., 2019 | 0.0001 | 0.0001 | 471 | 0.0001 | 0.0004 | 449 |
| Total (fixed effect, 95% CI) | | | 542 | | | 490 |
| Total (random effect, 95% CI) | | | | | | |

Heterogeneity: $Tau^2$ = 0.0454; $Chi^2$ = 3.98, df = 2 ($P$ = 0.1367); $I^2$ = 49.7%
Test for overall effect (fixed effect):Z= -3.06 ($P$= 0.0022)
Test for overall effect (random effects):Z= -1.94 ($P$= 0.0520)

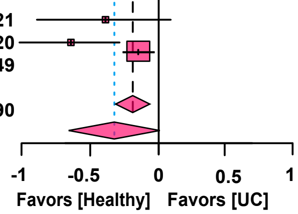


#### I. *Alistipes shahii*

| Study | UC Mean | UC SD | UC Total | Healthy Mean | Healthy SD | Healthy Total |
|---|---|---|---|---|---|---|
| Imhann F et al., 2016 | 0.0008 | 0.0023 | 23 | 0.0014 | 0.0019 | 21 |
| Morgan XC., 2012 | 0.0009 | 0.0016 | 48 | 0.0038 | 0.0029 | 20 |
| Total (fixed effect, 95% CI) | | | 71 | | | 41 |
| Total (random effect, 95% CI) | | | | | | |

Heterogeneity: $Tau^2$ = 0.5485; $Chi^2$ = 7.14, df = 1 ($P$ = 0.0076); $I^2$ = 86.0%
Test for overall effect (fixed effect):Z= -4.15 ($P$< 0.0001)
Test for overall effect (random effects):Z= -1.53 ($P$= 0.1270)

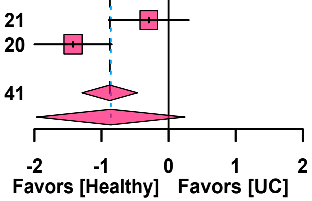


#### J. *Bacteroides eggerthii*

| Study | UC Mean | UC SD | UC Total | Healthy Mean | Healthy SD | Healthy Total |
|---|---|---|---|---|---|---|
| Imhann F et al., 2016 | 0.0025 | 0.0120 | 23 | 0.0017 | 0.0037 | 21 |
| Morgan XC., 2012 | 1.0e-9 | 1.0e-9 | 48 | 0.0223 | 0.0377 | 20 |
| Lloyd-Price J et al., 2019 | 0.0111 | 0.0413 | 471 | 0.0130 | 0.0405 | 449 |
| Hall AB et al., 2017 | 1.0e-9 | 1.0e-9 | 97 | 0.0038 | 0.0139 | 74 |
| Total (fixed effect, 95% CI) | | | 639 | | | 564 |
| Total (random effect, 95% CI) | | | | | | |

Heterogeneity: $Tau^2$ = 0.1846; $Chi^2$ = 16.97, df = 3 ($P$ = 0.0007); $I^2$ = 82.3%
Test for overall effect (fixed effect):Z= -2.36 ($P$= 0.0181)
Test for overall effect (random effects):Z= -1.44 ($P$= 0.1491)

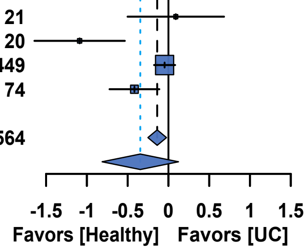
